# The Impact of Negative Emotions on Adolescents’ Nonsuicidal Self-Injury Thoughts: An Integrated Application of Machine Learning and Multilevel Logistic Models

**DOI:** 10.1101/2025.02.15.25322355

**Authors:** Chan-Young Ahn, Jin-Ha Kim, So-Jung Kim, Jae-Won Kim, Jung-Jo Na, Dong-Gi Seo, Jong-Sun Lee

## Abstract

Non-Suicidal Self-Injury (NSSI) is a prevalent and complex behavior among adolescents, often linked to negative emotions such as loneliness, anxiety, and emptiness. Traditional retrospective methodologies often fail to capture the dynamic and real-time nature of NSSI and its emotional triggers due to memory biases and temporal fluctuations. This study aimed to identify emotional predictors of NSSI thoughts among adolescents using machine learning and multilevel logistic regression.

The study included 42 adolescents (aged 12–15 years) who had engaged in NSSI in the past year. Participants reported their mood and NSSI behaviors three times daily over a 14-day EMA period via a smartphone application. Predictor variables included depression, anxiety, loneliness, self-anger, anger towards others, shame, and emptiness. A random forest model identified loneliness (feature importance: 0.40), anxiety (0.18), and emptiness (0.14) as the most significant predictors of NSSI thoughts. Multilevel logistic regression confirmed these findings, showing that each one-unit increase in anxiety, loneliness, and emptiness corresponded to a 24%, 19%, and 24% increase in the odds of experiencing NSSI thoughts, respectively. The ICC value of 0.26 indicated substantial between-individual variance, justifying multilevel modeling. However, random effects analysis revealed no significant individual differences, suggesting uniform effects across participants.

These findings highlight loneliness as the most influential predictor, emphasizing the need to address social connections in interventions. Combining machine learning with traditional statistical methods enhanced interpretability, providing practical insights for developing tailored, emotion-focused interventions for adolescents engaging in NSSI.

## Introduction

Nonsuicidal Self-Injury (NSSI) is defined as the deliberate and direct harming of one’s own body tissues without the intent to die (1). Specifically, cutting the body with a sharp instrument is a common attempt, and other self-injurious behaviors, such as burning or intentionally hitting oneself, occur in a variety of ways and intensities (2, 3). According to the diagnostic criteria proposed in the DSM-5 (Fifth Edition of the Diagnostic and Statistical Manual of Mental Disorders), NSSI applies to individuals who have engaged in deliberate self-harm without suicidal intent on 5 or more days in the past year (4). NSSI typically begins in early adolescence and may persist for several years (4). A meta-analysis study on the prevalence of NSSI (5) found that prevalence peaked in adolescence at 17.2%, followed by 13.4% in early adulthood and 5.5% in adulthood, with a gradual decline in prevalence after adolescence. Another study found that globally, the lifetime prevalence of self-injurious behavior among adolescents is 19% and 20% for non-suicidal self-injury (6). However, the actual prevalence of NSSI is likely to be higher than this, given that self-injurious behaviors are often secretive and private due to social stigma and concerns about being seen, and no specific medical attention is sought (7, 8).

Self-injurious behavior is highly associated with several psychiatric symptoms. Specifically, depressive and anxiety disorders and borderline personality disorder are highly associated with NSSI (2, 9, 10), as are eating disorders and posttraumatic stress disorder (2, 11, 12). It has also been found to be associated with a variety of psychiatric disorders, including attention deficit hyperactivity disorder, bipolar disorder, and substance use problems (9, 10, 13, 14). In particular, a serious problem with self-injurious behavior is its high association with suicide. Several studies have shown that, paradoxically, non-suicidal self-injury is a predictor of future suicidal behavior (15, 16), and actual suicide attempt intent reported in 70.8% of adolescents who engage in self-injury repeatedly (17). An individual’s ability to directly carry out a suicide is referred to as suicidal capability (18), and repeated self-injurious behaviors are quite dangerous in that they increase suicidal capability by decreasing fear of suicide, which ultimately increases the risk of suicide (18, 19). Repeated self-injurious behaviors during adolescence can make it difficult to succeed in school, career, and interpersonal relationships, which are key developmental tasks in early adulthood (20).

A growing body of research has identified factors that contribute to the development and maintenance of NSSI, aligning with several theoretical models (1, 3, 21). Collectively, the results suggest that one of the most common reasons for engaging in self-injurious behavior is to avoid or regulate negative emotions. Negative emotions that have been reported to play a role in NSSI include depression, anxiety, loneliness, anger toward self or others, rejection, and guilt (22–26). Individuals who report NSSI experience high levels of negative emotions (27), with some research utilizing longitudinal designs showing that negative emotions increase prior to the self-injurious behavior and decrease in the hours following the self-injurious behavior (12, 22). However, the alleviation of negative emotions caused by self-injurious behaviors is temporary and does not address the underlying issue, leading to the recurrence of such behaviors.

Despite these established connections between NSSI and negative emotions, understanding the timing, mechanisms, and motivations behind self-injury in daily life remains limited (28). The nature of NSSI and its associated emotions changes over time and across contexts, making it difficult to measure with one-time data collection. Traditional self-report and experimental studies relying on autobiographical memory are prone to biases, limiting ecological validity (29). Thus, meticulous observational studies are needed to measure temporal changes and comprehensively record diverse aspects of NSSI (30). Although some longitudinal studies identify developmental risk factors over extended periods (31), they focus on long-term trends at the between-person level, often lacking the temporal precision to capture the immediate risks of self-injury within short timeframes, such as minutes or hours in daily life. Ecological Momentary Assessment (EMA) repeatedly records individuals’ real-time experiences, allowing for the measurement of changes in behavior and understanding of situational effects with high ecological validity and minimal recall bias (32). Studies show that EMA is effective for exploring NSSI even with small samples (33, 34).

Studies exploring risk factors for NSSI using EMA have been thus increasing, but their predictive power remains weak (31, 35). This limitation appears to be partly due to an over-reliance on traditional statistical methodologies (35). Traditional approaches impose linearity on relationships that may have complex associations and limit the number of variables that can be examined simultaneously, forcing researchers to rely on overly simplified predictive models (36, 37). To address these issues, this study utilized machine learning techniques to iteratively test all possible relationships among variables and to construct predictive models that account for non-linear relationships.

In investigating NSSI, there is a growing trend towards actively utilizing machine learning approaches such as logistic regression and random forest (38, 39). According to a systematic review study, machine learning techniques have demonstrated higher predictive accuracy for self-injurious thoughts and behaviors (SITB) compared to traditional statistical methods (36). This suggests that machine learning methods can be reliably used for variable selection concerning SITB risk factors. Notably, machine learning techniques, such as random forest, offer advantages by identifying complex patterns and capturing non-linear relationships between variables. These methods can manage numerous predictors, providing feature importance scores that highlight key factors influencing outcomes. The use of machine learning for exploratory analysis can uncover hidden patterns and generate hypotheses, guiding further research and theory development (40, 41).

However, despite these strengths, such methods are often challenging to apply in confirmatory research due to their low interpretability. To address this limitation, this study complemented machine learning techniques with logistic regression analysis to enhance interpretability by allowing researchers to directly select and examine variables of interest (42). Finally, the aim of this study is to explore key emotional predictors of NSSI thoughts in adolescents by employing machine learning techniques alongside multilevel logistic regression analysis.

## Materials and Methods

### Participants

This study included 42 adolescents aged 12 to 15 years. The inclusion criteria were that both the adolescents and their parents voluntarily agreed to participate in the research, and the adolescents had engaged in NSSI on 1 or more days in the past year. Participants were recruited on a nationwide scale from multiple elementary and middle schools in South Korea.

Exclusion criteria for participants are as follows: presenting suicidal ideation and plans at a level requiring immediate intervention; current manifestation of psychotic or manic symptoms or substance use disorders; currently receiving any psychotherapy or counseling; and parental consent.

### Procedure

Participant recruitment for the study was conducted through DataSpring, a professional research firm, following predetermined eligibility criteria. The recruitment process involved two rounds of screening. Initial invitations were distributed to registered panel members (Round 1: 18,043 people, Round 2: 5,171), yielding 3,123 individuals responded to the screening questionnaire (Round 1: 2,225 people, Round 2: 898). Respondents received detailed information of the study, including its purpose, the procedures, potential risks, and benefits. Participants were informed of their right to withdraw at any time without consequences. Through this screening process, 111 eligible participants were identified (Round 1: 92 participants, Round 2: 19). Finally, 48 adolescents and their legal guardian (e.g., parents) finally agreed to participate in the study by selecting “yes’ to indicate their consent. As the study was conducted online, online informed consent was obtained instead of written informed consent, as approved by the Institutional Review Board (KWNUIRB-2022-01-009-002). Participant recruitment was conducted from May 25, 2022, to June 4, 2022.

The final sample in the analysis comprised 42 participants, after excluding one participant who withdrew during the study and five participants who failed to meet the minimum 50% ecological momentary assessment (EMA) response rate criterion. This systematic screening and enrollment process ensured the quality and reliability of the collected data while maintaining ethical standards for research with adolescent participants.

Upon the completion of the informed consent process, participants were integrated into the study through the utilization of the dedicated application, Mind Care, purposefully developed for this research endeavor. Subsequently, participants underwent a preassessment, involving the completion of a comprehensive questionnaire encompassing the Columbia Suicide Severity Rating Scale (C-SSRS), Center for Epidemiological Studies Depression Scale for Children (CES-DC), and Revised Children’s Manifest Anxiety Scale (RCMAS).

Throughout a 14-day EMA period, participants utilized a smartphone app to report their mood three times daily and document instances of NSSI behaviors and the intensity of NSSI thoughts. This resulted in a cumulative total of 41 assessments over the specified timeframe, with participants engaging in assessments three times a day for 13 days and twice a day on the concluding day.

Following the conclusion of the 14-day EMA period, participants were subjected to a postassessment utilizing the same questionnaire employed during the preassessment phase.

### Measures

#### Center for Epidemiological Studies Depression Scale for Children (CES-DC)

Developed by Weissman et al. (1980) (43), the Depression Scale for Children is a scale used to measure depression in adolescents. It consists of 20 items and measures depressive mood over the past week on a 4-point Likert scale (0: not at all, 3: very much). The Cronbach’s alpha in this study was .828.

#### Revised Children’s Manifest Anxiety Scale (RCMAS)

The Revised Children’s Manifest Anxiety Scale developed by Reynolds and Paget (44) was used to assess the level of anxiety in adolescents. The RCMAS is designed to measure the presence of various anxiety-related symptoms and is applicable to adolescents. It consists of 37 items with a 2-point Likert scale of “yes” (1 point) or “no” (0 point) for each item. The Cronbach’s alpha in this study was .823.

#### The Self-Harm Screening Inventory (SHSI)

The Self-Harm Screening Inventory (SHSI) is a brief self-report questionnaire designed to screen adolescents for self-harm behavior (45). It consists of 10 binary items (yes/no) that inquire about self-harm behavior over the past year. In this study, self-harm behavior was assessed using a 6-point Likert scale (0: Not applicable / 1: Once / 2: 2–5 times / 3: 6–10 times / 4: 11–30 times / 5: More than 31 times) to capture the frequency and types of self-harm, as well as to evaluate whether medical treatment was sought. The Cronbach’s alpha in this study was .916.

#### Columbia Suicide Severity Rating Scale (C-SSRS)

The Columbia Suicide Severity Rating Scale (C-SSRS) was developed by Posner, Brent (46) to assess the severity of and changes in suicidal ideation and behavior. It is designed to measure four components: the severity of suicidal ideation, the intensity of suicidal ideation, suicidal behavior, and lethality. Additionally, it accommodates different assessment periods based on research or clinical needs. In this study, one item assessing active suicidal ideation with some intent to act, without specific plan, and another item assessing active suicidal ideation with specific plan and intent were used to measure the presence or absence of suicidal intent and attempts over the past month, using dichotomous "yes" or "no" responses.

### Self-Injury and Suicidality Questionnaire

The Self-Injury and Suicidality Questionnaire is composed of binary items (yes/no) to assess the presence of self-injurious and suicidal thoughts, as well as self-injurious and suicidal behaviors, since the previous EMA session. If participants reported engaging in self-injurious behavior since the last assessment, additional questions were presented to inquire whether they experienced suicidal thoughts during or immediately before the self-injury, as well as what methods were used for self-injury.

### Mood Appraisal Questionnaire

Participants responded their emotions they are experiencing at that moment on a VAS 3 times a day. The mood Vas is consisted of a horizontal line from “not at all” at one end to “very much” at the other. Specifically, participants are instructed to respond to their feelings of depression, anxiety, shame, anger towards myself, anger towards others, loneliness, and emptiness on an 8-point Likert scale.

### Data analysis

This study aims to elucidate the causal factors behind thoughts of non-suicidal self-injury (NSSI). To achieve this, we employed a mixed-methods approach, focusing initially on quantitative analysis followed by a multilevel logistic regression to account for the nested structure of the data. The initial quantitative analysis was conducted using a random forest algorithm. Random forest, developed by Breiman (47), builds upon the decision tree classifier, which determines optimal split variables and points at each node using impurity measures such as chi-square statistics, Gini index, and entropy (48). While decision trees are more interpretable than other supervised learning techniques, they are often less predictive and can be highly sensitive to data variations (49). Random forest addresses these shortcomings by incorporating bootstrap aggregation and randomization of variables to enhance randomness and robustness. By randomly selecting subsets of variables, it generates diverse decision trees, reducing their correlation and improving prediction accuracy (50). This method was chosen for its robustness in handling a large number of predictor variables and for its utility in feature importance evaluation. The dependent variable in this analysis was the occurrence of NSSI thoughts, coded as a binary outcome (0 for absence, 1 for presence). Predictor variables included measures of depression, anxiety, loneliness, self-anger, anger towards others, shame, and emptiness.

In the random forest analysis, the model was configured with a maximum depth of three for each decision tree, ensuring that the model was sufficiently complex to capture relevant patterns in the data without overfitting. The ensemble consisted of 1,000 trees, providing a robust aggregation of decisions from multiple trees to improve the overall predictive accuracy and stability of the model. For model validation, a 5-fold cross-validation approach was employed.

Through the random forest analysis, we calculated the feature importance scores to identify which variables most significantly influenced the decision-making process in the algorithm. Feature importance in a random forest is determined based on how much each feature decreases the impurity of the split, which is typically measured by the Gini impurity in classification problems. This attribute provides a numerical value for each feature, representing its relative importance in the model (51).

A critical limitation of the random forest analysis was its inability to account for the multilevel structure of the data. Each participant provided multiple observations, creating a nested data format that the random forest model does not inherently consider. In social science research, an Intraclass Correlation Coefficient (ICC) value greater than 0.20 typically necessitates a multilevel analysis approach (52).

To address this limitation, we conducted a multilevel logistic regression analysis including fixed and random effects of predictors (53). The multilevel logistic regression, also known hierarchical logistic regression model, is particularly suited for analyzing data with a nested structure. This model helps capture the variability at both the within and between levels, offering a comprehensive understanding of how predictors at different levels influence the binary outcome. The dependent variable remained the binary indicator of NSSI thought occurrence. Given that our predictors are all related to negative emotional states and share considerable covariates, we carefully selected variables for inclusion in the regression model. Instead of including all predictors simultaneously, which could lead to multicollinearity issues, each predictor was included in separate models (54). This approach allowed us to isolate the effect of each predictor on NSSI thoughts while controlling for the nested data structure.

The analysis using Random Forest was conducted with Python version 3.10.12. To estimate the multilevel logistic regression model, Bayesian estimator in Mplus 8.3 was used.

## Results

### Demographic characteristics

Before conducting the main analysis, a descriptive statistical analysis was performed to examine the demographic characteristics of the participants. Among the participants, 64.3% were female (n=27) and 35.7% were male (n=15). The majority of participants were third-year middle school students (78.6%, n=33), followed by second-year middle school students (16.7%, n=7), with first-year middle school student (2.4%, n=1).

Regarding the participants’ self-perceived household economic status, the middle class category represented the largest proportion (45.2%, n=19), followed by upper-middle (33.3%, n=14), lower-middle (16.7%, n= 7), and low (4.8%, n=2). Regarding household size, four-person households were most common (45.2%, n=19), followed by three-person households (33.3%, n=14), five-person households (16.7%, n=7), and six-person households (4.8%, n=2).

### Random forest

The average EMA response rate was 86.5%. The confusion matrix, shown in Table 1, provides a comprehensive overview of the model’s performance. Using formulas 1.1 to 1.5, we derive the following evaluation metrics for the model: The accuracy of the model is 0.792, calculated as the ratio of true positives and true negatives to the total number of instances. However, relying solely on accuracy can be misleading when assessing the model’s performance, especially in the presence of class imbalance. Therefore, additional metrics are considered to provide a more detailed evaluation.

**Table 1.** Confusion Matrix for Binary Classification.

|  |  | Predicted |  |
| --- | --- | --- | --- |
|  |  | 0 | 1 |
| Actual | 0 | True Negative(TN)<br>810 | False Positive(FP)<br>303 |
|  | 1 | False Negative(FN)<br>189 | True Positive(TP)<br>114 |

The sensitivity, or recall, is 0.376, indicating that the model correctly identifies 37.6% of the actual positive cases. Specificity, which measures the model’s ability to correctly identify negative cases, is 0.728. The positive predictive value (PPV), representing the proportion of positive results that are true positives, is 0.273. Conversely, the negative predictive value (NPV), which measures the proportion of negative results that are true negatives, is relatively high at 0.811.

This data suggests that while the model performs well in identifying negative instances, its sensitivity is low due to the zero-inflated nature of the dataset, where instances of class 0 make up 21.3% of the total data. Consequently, the model exhibits a higher NPV because it accurately predicts the abundant negative instances. However, the PPV is low, reflecting the difficulty in correctly predicting positive instances. Despite the lower PPV of 0.273, this value is considered relatively high compared to other studies, indicating that the model performs reasonably well in identifying positive cases (28).

The feature importance of the random forest is viewed in Figure 1. The y-axis in Figure 1 represents the feature importance scores generated by the random forest model. These scores indicate the relative importance of each predictor variable in influencing the model’s predictions. Feature importance in a random forest is determined based on how much each feature decreases the impurity of the split, which in this case is measured using the Gini impurity for classification problems. Among all variables, loneliness had the highest score of 0.40. Anxiety ranked as the second highest among all variables with a score of 0.18, followed by emptiness, which had a score of 0.14. Depression, self-anger, anger towards others, and shame showed similar levels of feature importance scores in the analysis (Depression = 0.08, self-anger = 0.07, anger towards others = 0.07, shame = 0.07).

**Figure 1.**
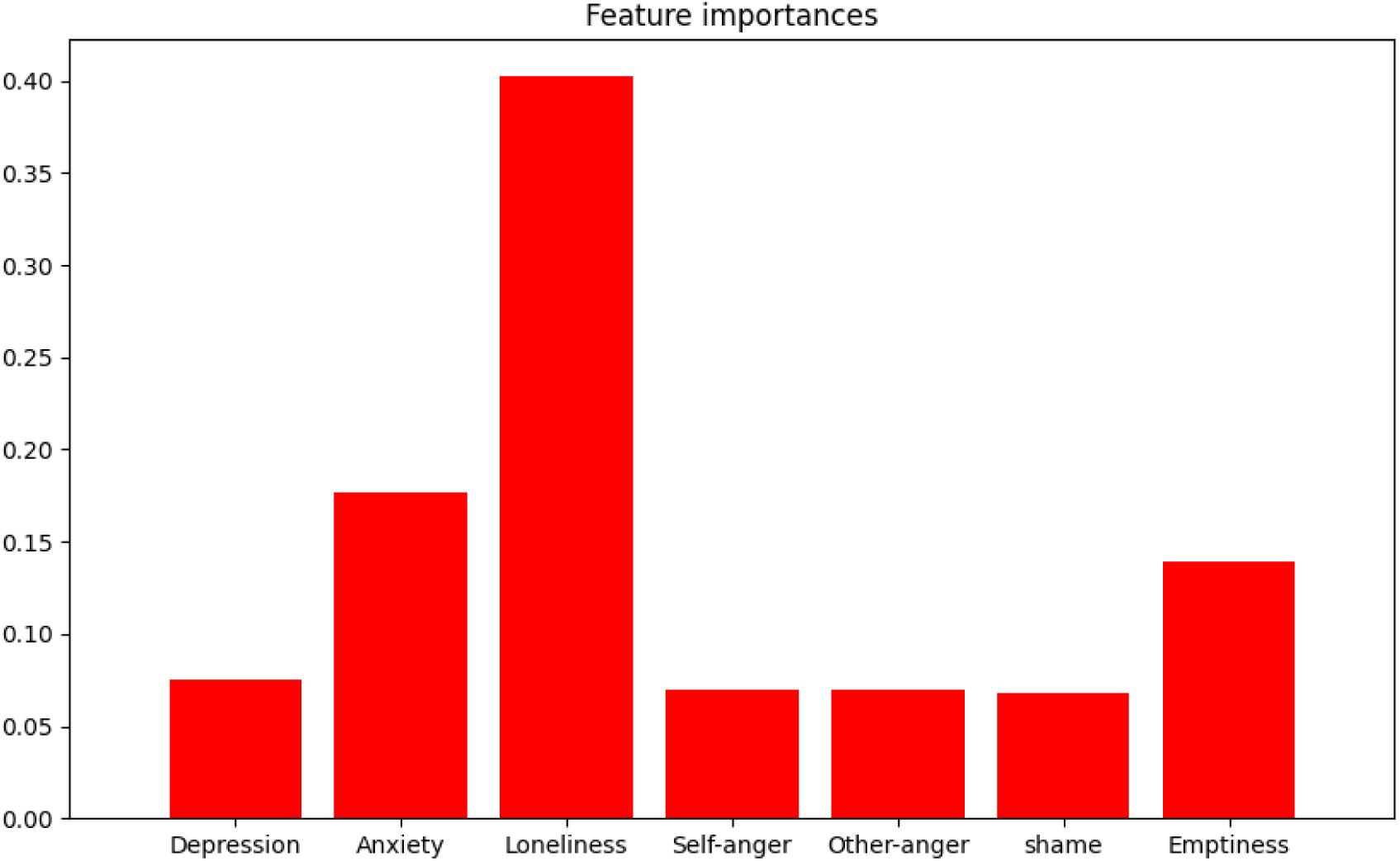
Feature importance scores of the predictors.

### Multilevel logistic regression

The ICC(Intraclass Correlation Coefficient) value for the dependent variable ‘occurrence of NSSI thoughts’ collected in the research was found to be 0.26. An ICC value of 0.26 indicated that the between level variance accounted for approximately 26% of the total variance in the dependent variable. This suggests the need for employing a multilevel model in analyzing this data. Table 2 displays the result of a multilevel logistic regression model that predicts NSSI thought with depression, anxiety, loneliness, self-anger, anger towards others, shame, and emptiness as predictor variables. The detailed equations of the multilevel logistic regression models are presented in Appendix 1. In the fixed effects analysis, the variables anxiety, loneliness, and emptiness were found to be significant [anxiety (0.216, *p* = 0.005), loneliness(0.177, *p* = 0.013), emptiness(0.217, *p* = 0.027)]. For each one-unit increase in anxiety, there is a 24% increase in the odds of experiencing an NSSI thoughts [exp(.216) = 1.24]. For each additional unit increase in loneliness, the odds of NSSI thoughts increase by 19% [exp(.177) = 1.19]. An increase in the measure of emptiness by one unit is associated with a 24% increase in the odds of experiencing NSSI thoughts [exp(.217) = 1.24]. It is noteworthy that the outcomes of the random forest’s feature importance scores and the multilevel logistic regression analysis are in concordance. In the random effects analysis, none of the variables were found to be significant. This suggests that there were no individual differences in the impact of these variables on NSSI thoughts, indicating a uniform influence of these variables across individuals.

**Table 2.** Multilevel logistic regression models with each negative emotions as a predictor.

|  | Effects | Estimate | S.E | p-value |
| --- | --- | --- | --- | --- |
| Depression | Fixed effect($\gamma_{00}$) | 0.196 | 0.127 | .123 |
| | Random effect($\tau_{00}$) | 0.044 | 0.062 | .475 |
| Anxiety | Fixed effect($\gamma_{10}$) | 0.216 | 0.077 | .005* |
| | Random effect($\tau_{10}$) | 0.000 | 0.000 | .634 |
| Loneliness | Fixed effect($\gamma_{20}$) | 0.177 | 0.071 | .013* |
| | Random effect($\tau_{20}$) | 0.000 | 0.000 | .500 |
| Self-anger | Fixed effect($\gamma_{30}$) | 0.117 | 0.149 | .432 |
| | Random effect($\tau_{30}$) | 0.040 | 0.048 | .404 |
| Anger towards others | Fixed effect($\gamma_{40}$) | -0.056 | 0.168 | .738 |
| | Random effect($\tau_{40}$) | 0.198 | 0.134 | .141 |
| Shame | Fixed effect($\gamma_{50}$) | -0.270 | 0.392 | .490 |
| | Random effect($\tau_{50}$) | 0.463 | 0.683 | .498 |
| Emptiness | Fixed effect($\gamma_{60}$) | 0.217 | 0.099 | .027* |
| | Random effect($\tau_{60}$) | 0.001 | 0.011 | .921 |

Note. An asterisk (\*) indicates that the p-value is less than .05.

## Discussion

The present study aimed to identify negative emotions that predict NSSI thoughts using random forest techniques and multilevel logistic regression. The key findings show that loneliness emerged as the most influential variable, with a feature importance score of 0.40, followed by anxiety and emptiness. These results were also confirmed by multilevel logistic results. These findings underscore the substantial impact of loneliness on the occurrence of non-suicidal self-injury thoughts in adolescents, suggesting that interventions aimed at reducing loneliness could be a key focus when addressing self-injurious thoughts within this group. This aligns with previous research findings that suggest loneliness is a significant factor influencing non-suicidal self-injury in adolescents (26, 55).

Adolescence is a critical period marked by evolving social connections and a heightened need for intimacy, factors that contribute to the persistence of loneliness (56). This stage is crucial for understanding the dynamics between loneliness and Non-Suicidal Self-Injury(NSSI), which can be conceptualized within the interpersonal functioning model of NSSI (1). During adolescence, experiences such as social distress from bullying and/or academic pressure from examinations are significant stressors that may lead adolescents to engage in NSSI. These behaviors serve not only as a means to alleviate emotional distress but also play a complex role in social interactions (57). Specifically, NSSI functions as a cry for help or a method to gain support, thereby strengthening social bonds. This social reinforcement can inadvertently maintain and even reinforce the cycle of self-injurious behavior, as suggested by Nock and Prinstein (58).

Furthermore, according to the interpersonal theory of suicide, thwarted belongingness, a feeling stemming from loneliness or isolation, substantially elevates an individual’s risk of suicide (18, 59). Indeed, the interplay between NSSI and suicidal thoughts and behaviors has been documented, showing a close associations, which suggests that underlying interpersonal states fostered by loneliness are critical contributors to both NSSI and suicidal behaviors (60).

Given these insights, it is imperative for clinicians to design targeted interventions that address loneliness when developing strategies for adolescents engaging in self-injury. Such interventions could focus on enhancing social connectedness by fostering positive and supportive relationships with family, friends, and peers. In addition, efforts may be necessary to disseminate and promote psychoeducational manuals on emotional regulation strategies aimed at reducing loneliness in self-injuring adolescents, targeting teachers and clinicians as key disseminators (61).

Our study shows emptiness is one of the important factors we need to note in accounting for adolescents’ self-harm. While research examining the direct relationship between emptiness and NSSI has been limited, existing evidence suggests that emptiness may lead individuals to engage in NSSI as a means to enhance emotional and sensory experiences (62). Prior research has demonstrated that emptiness is significantly associated with suicidal ideation and attempts (63). In a study of women diagnosed with borderline personality disorder (BPD), feelings of emptiness preceded NSSI, suggesting that NSSI may serve as a coping mechanism for negative emotional states, particularly emptiness (64). Furthermore, research involving college students who engaged in NSSI, 67% reported that feelings of emptiness sometimes preceded their NSSI behaviors (63). Other studies involving BPD populations have shown that, compared to impulsivity, affective instability, and anger, emptiness is the only BPD symptom that predicts all eight indicators of psychosocial morbidity, including suicidality (65). Moreover, individuals who experience maltreatment during childhood are at increased risk of experiencing emptiness during adolescence and adulthood. This psychological condition is theorized to increase the likelihood of engaging in NSSI as a means of alleviating feelings of emptiness and achieving a sense of well-being (66, 67). The present findings are align with and extend these previous studies.

NSSI is associated with anxiety, which is considered a significant trigger for these behaviors (68, 69). For example, NSSI is often utilized as a method to alleviate distress related to anxiety symptoms (70), and anxiety reduction is frequently reported as one of the primary functions of NSSI (71). Additionally, individuals who meet the diagnostic criteria for an anxiety disorder—characterized by frequent and intense negative emotions and efforts to avoid or escape such experiences—report higher rates of NSSI compared to those without an anxiety disorder diagnosis (72). Furthermore, a positive association between anxiety and NSSI has also been observed in nonclinical community samples (73). Consistent with these previous findings, our results identified anxiety as a key factor of NSSI among adolescents, further supporting the crucial role of anxiety in understanding and addressing adolescents’ self-harming behaviors.

The relationship between NSSI and anxiety can be understood within the framework of one of the widely accepted theories, the emotional cascade theory (74). This theory suggests that even minor emotional stimuli can trigger repetitive negative thinking (e.g., rumination), which amplifies the experience of negative emotions and leads to a vicious cycle, or an emotional cascade (75). In an attempt to halt this emotional cascade, individuals may resort to extreme forms of distraction, such as NSSI. Following this, they often experience a sense of relief due to negative reinforcement, which is analogous to the relief experienced after avoidance in the context of anxiety.

The Intraclass Correlation Coefficient (ICC) value of 0.26 in the present study indicates that 26% of the variance in NSSI thoughts can be attributed to between-individual differences. This justified the utilization of a multilevel model, allowing for a nuanced examination of the impact of various predictors. The results of the fixed effects analysis highlighted the significance of anxiety, loneliness, and emptiness. Each unit increase in anxiety, loneliness, and emptiness corresponded to a 24%, 19%, and 24% increase in the odds of experiencing NSSI thoughts, respectively. These findings align with the random forest results, reinforcing the robustness and consistency of the identified predictors.

The noteworthy concordance between the random forest’s feature importance scores and the results of the multilevel logistic regression analysis enhances the reliability of the identified predictors. Both approaches underscored the pivotal role of loneliness, anxiety, and emptiness in influencing NSSI thoughts. This convergence strengthens the validity of the study’s findings, demonstrating a consistent pattern across different analytical methodologies. Furthermore, this suggests that machine learning techniques such as random forest can be reliably applied to explore predictive factors that explain the characteristics of non-suicidal self-injury (36, 41).

In contrast, the random effects analysis revealed no significant individual differences, suggesting a uniform impact of the identified variables. This finding implies that, while the factors influencing NSSI thoughts are consistent, their intensity may vary among individuals. The absence of significant random effects highlights the universal relevance of loneliness, anxiety, and emptiness in shaping NSSI thoughts, irrespective of individual differences.

The strength of this study lies in its ability to capture fluctuations in emotions and self-injurious thoughts experienced by individuals in their daily lives through the use of digital-based methodologies. Variables related to NSSI, including emotions, exhibit fluctuations within a day (76). Therefore, traditional retrospective self-report methods may fail to adequately capture the emotional dynamics that change within minutes or hours in daily life. Thus, researchers need to devise meticulous and refined approaches for longitudinal and real-time data collection to assess self-injurious behavior and emotions (30).

Another strength of this study is the use of sophisticated statistical methodologies. In psychological research, particularly in studies of complex behaviors like NSSI, traditional statistical methods often face limitations due to the intricate interplay of numerous predictors. In our study, the integration of random forest analysis with multilevel logistic regression allowed us to robustly identify key predictors of NSSI thoughts, underscoring the potential of these advanced analytical approaches for gaining deeper insights into psychological phenomena and improving mental health outcomes.

While multilevel logistic regression analysis yielded non-significant random effects, suggesting a uniform influence of predictors across individuals, it is important to consider several limitations and future directions. The current sample size may have limited our ability to detect significant random effects. Future studies should aim for larger, more diverse samples to increase statistical power and generalizability.

A short observation period (14 days) limits our ability to examine how the relationship between predictors and NSSI thoughts may change over time. The use of a longer observation windows of longitudinal follow-up studies could provide insights into stability or variability of these relationships across different life stages or circumstances. While we found no significant random effects across the entire sample, there may be meaningful differences among subgroups. Future research could investigate potential moderators such as age, gender, or clinical diagnosis.

On the other hand, the growing interest in machine learning within psychology has led to the application of techniques like random forest in various research domains (77, 78). While these methods offer substantial advantages, such as the ability to manage large datasets and identify complex, non-linear relationships, they also present notable challenges. Specifically, random forest models suffer from low interpretability, making them difficult to use in a confirmatory context, particularly in social science research where understanding the relationship between variables is crucial. This limitation is especially pertinent in psychology, where the ability to interpret and communicate findings is essential (48, 79). Therefore, it is important to complement machine learning methods with traditional statistical techniques, such as logistic regression, where researchers can directly select and examine variables of interest. These classical methods not only enhance interpretability but also allow for hypothesis-driven confirmatory analysis, ensuring that research findings are both robust and theoretically sound (42).

## Conclusions

This study’s findings, derived from both random forest analysis and multilevel logistic regression, consistently identify loneliness, anxiety, and emptiness as key predictors of NSSI thoughts. While machine learning methods proved valuable for exploratory analysis, their integration with traditional statistical approaches enhanced the robustness of our results. The findings have important clinical implications, suggesting that NSSI interventions should be tailored to address the unique negative emotions of individuals with NSSI, rather than applying a one-size-fits-all approach. This personalized approach, focusing on each individual’s unique combination of negative emotions, may lead to more effective treatment outcomes. This research not only validates the complementary use of different analytical methods in psychological research but also provides practical insights for developing targeted NSSI interventions.

## Data Availability

The data underlying the findings described in this manuscript are not publicly available at this time. However, we are working with the journal to determine an appropriate method for data sharing. We will ensure that data access details are provided before publication.

## Acknowledgements

This work was supported by the Ministry of Health and Welfare of the Republic of Korea (HL19C00351).

## Appendix

### Appendix 1 Within-level

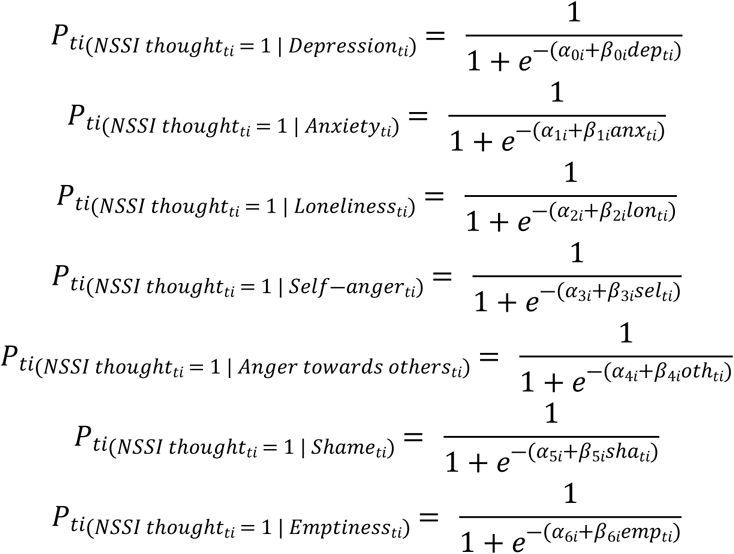

*NSSI thought_ti_* is the occurrence of the NSSI thought for the *t*-th report in *i*-th participant. The subscript *ti* for the predictors also represents the values of the predictors for the t-th report in the i-th participant.

### Between-level

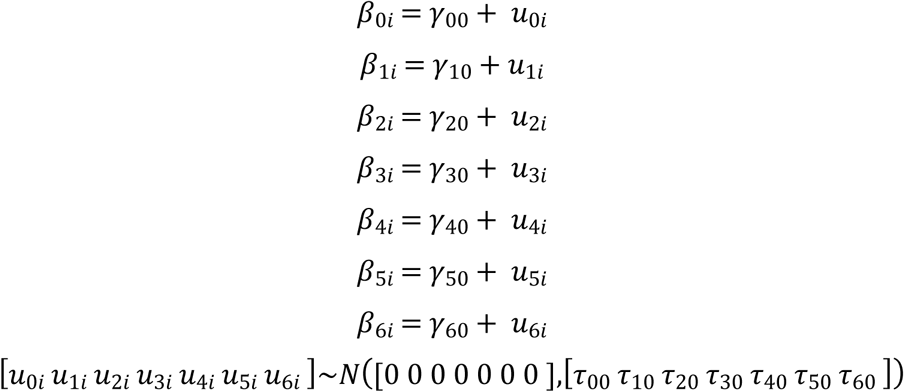

The subscript ’i’ in the between-level variables represents the slope of the i-th person for each variable. *γ*_00_, *γ*_01_, *γ*_02_, *γ*_03_, *γ*_04_, *γ*_05_, *γ*_06,_ are the fixed effect of the predictors. The vector of random effects, *u*_0*i*_, *u*_1*i*_, *u*_2*i*_, *u*_3*i*_, *u*_4*i*_, *u*_5*i*_, *u*_6*i*,_ are assumed to follow multivariate normal distribution with mean vector 0 and variance *τ*_00_, *τ*_01_, *τ*_02_, *τ*_03_, *τ*_04_, *τ*_05_, *τ*_06_

